# Seeing Beyond the Symptoms: Biomarkers and Brain Regions Linked to Cognitive Decline in Alzheimer’s Disease

**DOI:** 10.1101/2023.04.19.23288823

**Authors:** Seyed Hani Hojjati, Abbas Babajani-Feremi, the Alzheimer’s Disease Neuroimaging Initiative

## Abstract

**Objective:** Alzheimer’s disease (AD) is the most prevalent form of dementia, and its pathological process can only be detected through clinical approaches. Early diagnosis of AD is difficult, as most individuals with AD are not diagnosed in the early stages, and symptoms become more prominent as the disease progresses. Therefore, identifying specific biomarkers and predicting AD in the early stages is crucial. In this study, we aimed to identify effective biomarkers and brain regions that are strongly associated with cognitive decline in AD.

**Methods:** We included a large sample of 1759 individuals, covering a range of cognitive aging, including healthy controls (HC), mild cognitive impairment (MCI), and AD. We extracted nine different biomarkers based on three neuroimaging modalities: structural magnetic resonance imaging (sMRI), positron emission tomography (PET), and diffusion tensor imaging (DTI) to predict three neuropsychological scores: Mini-Mental State Examination (MMSE), Clinical Dementia Rating Scale Sum of Boxes (CDRSB), and Alzheimer’s Disease Assessment Scale-Cognitive Subscale (ADAS). Rather than integrating biomarkers, we monitored and explored the complex interrelated mechanisms underlying the development of AD separately for each biomarker. For prediction tasks, we used the ensemble regression tree by implementing bagging and random forest techniques in four different combination groups consisting of different subsets of subject categories (i.e., HC, MCI, and AD).

**Results:** Our results demonstrated that different biomarkers could predict all three cognitive scores, and we identified the most associated features with the cognitive scores, including (a) the right transverse temporal based on Amyloid-β (Aβ) deposition, (b) the left and right entorhinal cortex, left inferior temporal gyrus, and left middle temporal gyrus based on average cortical thickness (ATH), and (c) the left uncinate fasciculus based on mean diffusivity (MD).

**Conclusions:** The results of this study emphasize the significance of taking an interdisciplinary approach in comprehending the underlying mechanisms of AD. Additionally, these findings shed light on the diversity of the disease and have the potential to contribute to the development of more efficient treatments.

## 1. Introduction

Alzheimer’s disease (AD) is a condition that gradually affects brain function over time. It is a complex disease that results in progressive pathological changes in the brain’s biochemical and biological processes, leading to permanent impairment of cognitive functions [1–3]. Neuropsychological assessments are the initial and crucial step in characterizing symptoms of AD and evaluating diverse cognitive domains such as memory, language, and executive function [4–8]. However, diagnosing AD in its early stages is challenging as symptoms may not be fully present. Neuroimaging is an indispensable tool in enhancing early diagnosis, as changes in the brain occur years before symptoms appear [9–11]. Therefore, predicting neuropsychological assessments based on neuroimaging biomarkers in the early/asymptomatic and late/symptomatic stages of AD is crucial to understanding the presenting symptoms and identifying the relationship between different patterns of impairment in disease-related brain regions [12–14].

Neuroimaging can be highly effective in examining the progression of AD and identifying sensitive indicators in the early stages of the disorder. Various neuroimaging tools, including structural magnetic resonance imaging (sMRI), positron emission tomography (PET), and diffusion tensor imaging (DTI), have been used to predict AD progression through neuropsychological assessment [15, 16]. Recent studies have used volumetric biomarkers based on sMRI, such as the volume of gray matter and cortical thickness, to investigate the relationship between neuropsychological scores and neuroimaging biomarkers [12, 17–19]. The advent of PET has also enabled accurate investigation of the associations between neuropsychological assessment and proteinopathies during AD pathogenesis. While 18F-fluorodeoxyglucose (FDG), tau tangles, and Amyloid-β (Aβ) plaques are the key pathological markers of AD, previous studies have shown that Aβ deposition is weakly related to cognition [20]. In contrast, tau and FDG pathological changes have been reported as strong biomarkers that associate with cognitive decline [21–23]. Neuropathological studies suggest that tau mediates the relationship between Aβ and cognitive decline, which mostly emerges in patients with mild cognitive impairment (MCI) and AD [15, 24]. The amyloid cascade hypothesis suggests that the accumulation of Aβ initiates a series of events that ultimately lead to the development of AD. Specifically, it is believed that the accumulation of Aβ triggers inflammation and oxidative stress, which damages neurons and disrupts their communication [26, 25]. This, in turn, leads to the development of classic symptoms of AD, including memory loss, cognitive decline, and behavioral changes [27].

Recent studies have also utilized DTI for detecting micro-structural changes that are usually invisible in anatomical scans and would not be detected by PET [16, 28]. Compared with volumetric measures from sMRI and pathological biomarkers from PET imaging, DTI biomarkers are rarely used to predict neuropsychological assessments. Nevertheless, recent advancements in the neurobiology of AD suggest that AD is a multifactorial and heterogeneous disease that cannot be explained based on a single biomarker and a single modality [29]. Therefore, multimodal imaging techniques can help explore the complex, consistent changes accompanying AD.

The application of machine learning and regression analysis can aid in the early diagnosis and treatment of cognitive impairments, including AD. In recent years, various regression methods, such as least squares, support vectors, lassos, and regression trees, have been successfully used to predict neuropsychological scores based on neuroimaging biomarkers [15, 30, 31]. Machine learning has emerged as the most appealing technique for predicting cognitive scores [15, 32, 33]. Recent studies have also shown a strong interest in integrating features from different neuroimaging modalities to predict neuropsychological assessments based on machine learning techniques [15, 34]. Some studies have combined neuropsychological scores with neuroimaging biomarkers to find the progression trend or predict neuropsychological assessments [15, 35, 36]. However, the use of numerous predictors in such studies can lead to the inclusion of unrelated information in their different prediction tasks, leading to a decrease in regression performance [37]. Additionally, the use of cognitive assessments for predicting other cognitive assessments can result in bias due to their high correlation [15]. In recent years, neuropsychological predictions have commonly incorporated single-modal data or found integrative methods for combining data across multiple biomarkers based on multimodal data. However, different biomarkers across modalities have not adequately captured the heterogeneity of AD progression.

To our knowledge, prior research has largely focused on achieving high accuracy in classifying subjects or minimizing errors in estimating cognitive scores through regression analysis. However, most studies that have used multimodal feature domains have given little consideration to the differences between feature domains in sample data. To advance clinical and drug development research, it is crucial to pay more attention to the association between effective biomarkers and brain regions affected by AD during its progression stages. Therefore, the goal of this study is to bridge the gap between these research domains, providing significant value in the context of clinical and therapeutic investigations.

To achieve our objective, we analyzed different modalities and biomarkers separately instead of integrating them, exploring the progression of AD. We identified the most significant predictive biomarkers and brain regions for each cognitive assessment, utilizing three neuroimaging modalities (sMRI, DTI, and PET) to predict AD progression using the Mini-Mental State Examination (MMSE), Clinical Dementia Rating Sum of Boxes (CDRSB), and Alzheimer’s Disease Assessment Scale (ADAS). From a large sample set of 1759 individuals, covering a spectrum of cognitive aging, MCI, and AD, we extracted nine competing biomarkers.

Using the ensemble regression tree (ERT), we predicted target cognitive scores for four different combinations of subjects in various stages: healthy controls (HC)/MCI, HC/AD, MCI/AD, and HC/MCI/AD. By leveraging large sample sets, three complementary neuroimaging modalities, and nine different biomarkers, we were able to determine the extent to which each biomarker contributes to cognitive decline. To gain insights into AD progression, particularly in the early stages, we investigated the spatial and pseudo-temporal characteristics of neuroimaging biomarkers and their association with cognitive assessments. By developing a reliable biomarker based on each neuroimaging modality, we have the potential to capture the heterogeneity in the clinical evaluation of at-risk individuals and accelerate preventive strategies in the early stages of cognitive decline.

## 2. Materials and Methods

### 2.1. Participants

This study utilized MRI, DTI, PET, and neuropsychological data from a total of 1759 participants. The data subset was obtained from the Alzheimer’s disease prediction of longitudinal evolution (TADPOLE) challenge (https://tadpole.grand-challenge.org) [38], which was sourced from the Alzheimer’s Disease Neuroimaging Initiative (ADNI) database (adni.loni.usc.edu). The current study included 648 healthy controls (294 males), 699 individuals with MCI (411 males), and 412 individuals with AD (234 males). Table 1 presents the demographic and clinical characteristics of the participants. Unstable HC and MCI subjects with any conversions or revisions were excluded [39]. It is important to note that subjects did not necessarily require all three neuroimaging modalities, as each modality had been used in separate prediction tasks.

**Table 1.**
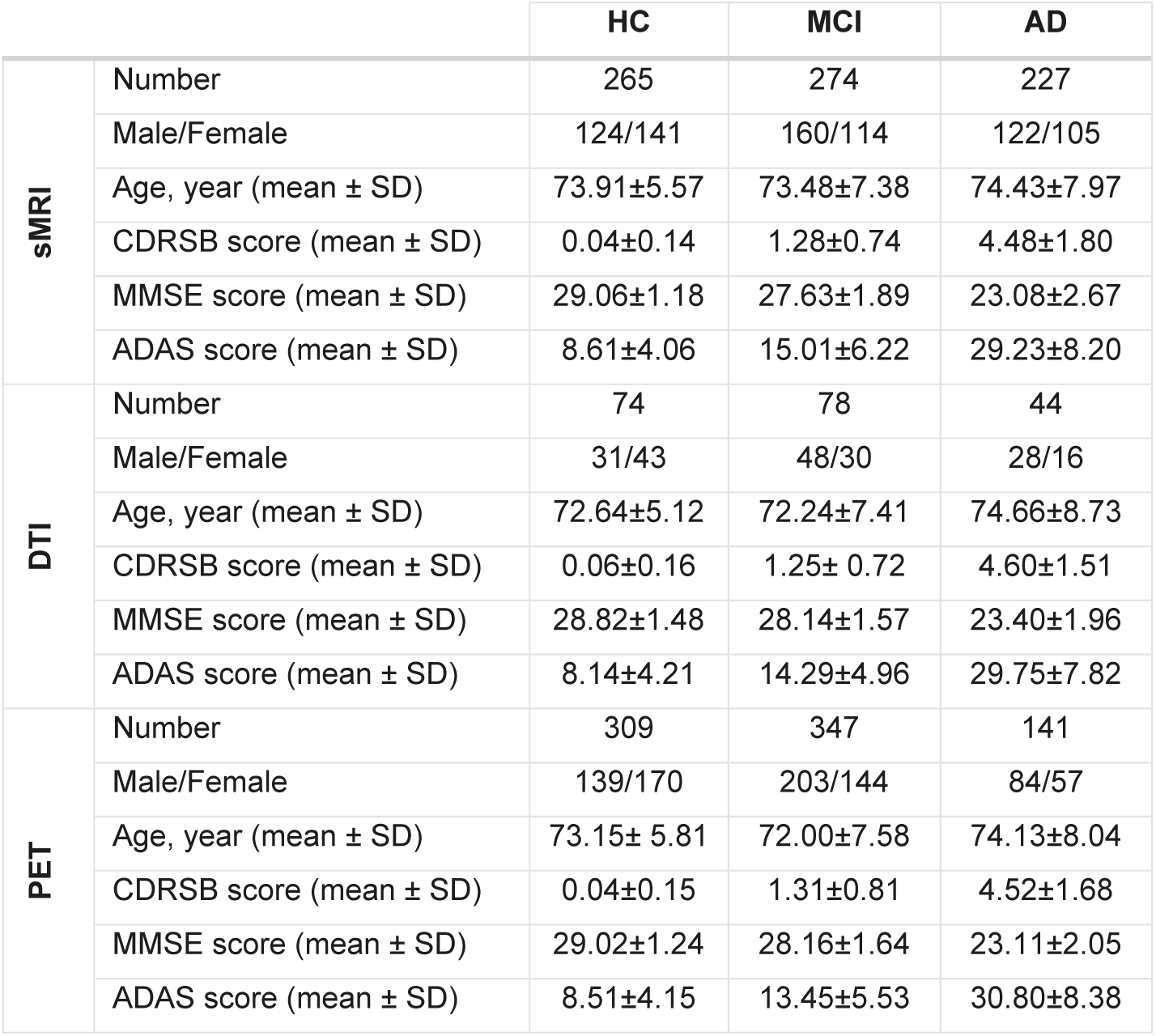
Demographic and clinical data.

|  |  | HC | MCI | AD |
| --- | --- | --- | --- | --- |
| sMRI | Number | 265 | 274 | 227 |
|  | Male/Female | 124/141 | 160/114 | 122/105 |
| | Age, year (mean ± SD) | 73.91±5.57 | 73.48±7.38 | 74.43±7.97 |
| | CDRSB score (mean ± SD) | 0.04±0.14 | 1.28±0.74 | 4.48±1.80 |
| | MMSE score (mean ± SD) | 29.06±1.18 | 27.63±1.89 | 23.08±2.67 |
| | ADAS score (mean ± SD) | 8.61±4.06 | 15.01±6.22 | 29.23±8.20 |
| DTI | Number | 74 | 78 | 44 |
|  | Male/Female | 31/43 | 48/30 | 28/16 |
| | Age, year (mean ± SD) | 72.64±5.12 | 72.24±7.41 | 74.66±8.73 |
| | CDRSB score (mean ± SD) | 0.06±0.16 | 1.25± 0.72 | 4.60±1.51 |
| | MMSE score (mean ± SD) | 28.82±1.48 | 28.14±1.57 | 23.40±1.96 |
| | ADAS score (mean ± SD) | 8.14±4.21 | 14.29±4.96 | 29.75±7.82 |
| PET | Number | 309 | 347 | 141 |
|  | Male/Female | 139/170 | 203/144 | 84/57 |
| | Age, year (mean ± SD) | 73.15± 5.81 | 72.00±7.58 | 74.13±8.04 |
| | CDRSB score (mean ± SD) | 0.04±0.15 | 1.31±0.81 | 4.52±1.68 |
| | MMSE score (mean ± SD) | 29.02±1.24 | 28.16±1.64 | 23.11±2.05 |
| | ADAS score (mean ± SD) | 8.51±4.15 | 13.45±5.53 | 30.80±8.38 |

### 2.2. Cognitive assessments

This study aimed to predict cognitive scores using imaging biomarkers. Three cognitive scores were used as targets for prediction: the MMSE, CDRSB, and ADAS. Each score highlights a different aspect of an individual’s neuropsychological status, depending on their stage of AD progression. The MMSE test is scored on a scale of 0 to 30, with a higher score indicating better cognitive function. The CDRSB and ADAS are scored on scales ranging from 0 to 18 and 0 to 70, respectively, with higher scores indicating more severe dementia and lower scores indicating milder dementia.

### 2.3. Neuroimaging data

We used various biomarkers from neuroimaging data as predictors. Nine different types of biomarkers were determined using a total of three neuroimaging modalities: sMRI, DTI, and PET. The aim of selecting these biomarkers was to implement a reliable machine learning approach based on a sufficient number of samples. All imaging data was already processed, and post-processed measurements for each biomarker were collected [38]. The sMRI biomarkers can be categorized into four groups: volume of gray matter (VGM) in 68 cortical brain regions, average thickness (ATH) in 68 cortical brain regions, surface region (SA) in 70 cortical and subcortical brain regions, and volume of white matter (VWM) in 45 brain regions. The DTI biomarkers consist of fractional anisotropy (FA), mean diffusivity (MD), radial diffusivity (RD), and longitudinal diffusivity (LD), with 57 brain regions each. 18F-florbetapir PET imaging was used to extract the Amyloid-β (Aβ) deposition in 109 cortical and subcortical brain regions.

### 2.4. Modeling the ERT

In this study, the ERT was used to predict cognitive scores from neuroimaging biomarkers. Firstly, the subjects were randomly divided into 500 training (90%) and test (10%) sets in each of the four combinations (HC/MCI, HC/AD, MCI/AD, and HC/MCI/AD). To reduce the bias and variance effect, ensemble algorithms were used to combine the predictions of several estimators [40]. This study employed bagging tree (BT) and random forest (RF) ensemble learning. A BT aggregation reduces the variance of a decision tree by grouping several weak learners into a strong one [41]. To create the BT algorithm, we trained 50 learners’ decision trees by randomly selecting subsets of data from training samples. The BT algorithm resulted in an ensemble (aggregation) of different models. In addition to growing trees, RF uses a random selection of features instead of applying all features at once. Random feature selection resulted in more independent learners, which led to better predictive performance due to the tradeoff between variance and bias. This study used an interaction test to split the RF predictors [42]. Predictors were split according to their minimum p-values from chi-square tests of independence. The importance of each feature was calculated as part of the RF analysis for each of the features used in our model. Trees were grown with surrogate splits to calculate the error function for a variable at each split point [43]. The predictor importance was estimated by summing the square errors at each branch node and dividing by the number of branches. The reported performances and predictor importance were calculated based on the average values across 500 iterations. In addition, t-tests were used to determine the significance of each biomarker and all of the reported mean squared error (MSE) was based on the test sets. The ERT algorithm and statistical analyses were implemented in MATLAB (2019b, Mathworks, Natick, MA).

### 2.5. Ranking the Significant Brain Regions

We conducted permutation tests 500 times under the null hypothesis to test our algorithm for ranking brain regions regarding their importance in the prediction of different stages of AD. To detect non-significant and unrelated features (i.e., brain regions), we performed the permutation test on combinations of HC, MCI, and AD by randomly shuffling the target scores (i.e., cognitive scores) for 500 iterations. We then used the mean plus two standard deviations (STD) of the permutation test results across features for each biomarker as a threshold to identify the most significant biomarkers for predicting cognitive scores. Finally, we arrived at a consensus of top-ranked features (brain regions) among the three prediction models based on comparative cognitive scores.

## 3. RESULTS

### 3.1. Prediction of Cognitive Scores Across Biomarkers and Combinations of Subjects

**Figures 1A-C** illustrate the prediction performances of the three cognitive scores (MMSE, CDRSB, and ADAS) across four different combinations of subjects (HC/MCI, HC/AD, MCI/AD, and HC/MCI/AD). A total of nine ERTs were trained to predict cognitive scores for each biomarker. According to **Figure 1A**, all nine biomarkers achieved comparable MSE values for predicting MMSE scores in HC/MCI combinations. However, when the AD category was present, Aβ outperformed the other eight biomarkers in prediction performance with MSE values of 4.47, 4.80, and 3.86 for HC/AD, MCI/AD, and HC/MCI/AD, respectively (p-value < 0.01). **Figures 1B and 1C** demonstrate that Aβ achieved similar results in CDRSB and ADAS scores. In these figures, Aβ provided the best prediction MSE values for CDRSB score with MSE values of 2.12, 1.96, and 1.69 for HC/AD, MCI/AD, and HC/MCI/AD, respectively (p-value < 0.01). Moreover, it provided the best prediction of MSE values of 60.94, 52.79, and 47.83 for ADAS for HC/AD, MCI/AD, and HC/MCI/AD, respectively (p-value < 0.01). The results showed a high association between cognitive scores and Aβ biomarkers in combinations where AD was present. Among sMRI biomarkers, ATH had the lowest MSE in all four combinations (e.g., HC/MCI), which was associated with more cognitive scores than the other three sMRI biomarkers. Considering the DTI biomarkers, MD achieved the least MSE in all four combinations.

**Figure 1.**
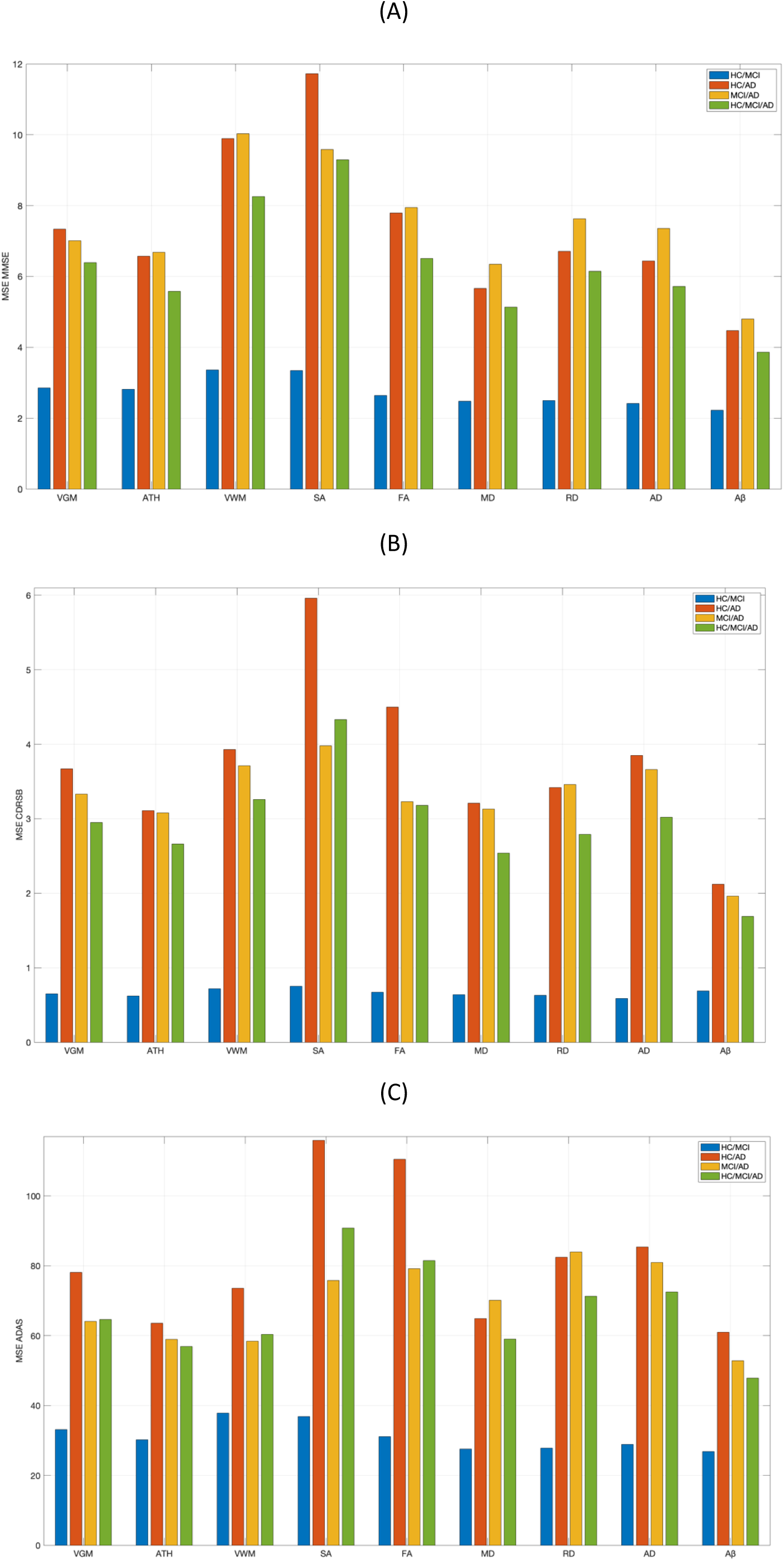
A) Mean squared error (MSE) of MMSE score. B) MSE of CDRSB score. C) MSE of ADAS prediction based on nine different biomarkers: VGM, ATH, VWM, SA, FA, MD, RD, LD, Aβ in four different combinations of groups of subjects: HC/MCI, HC/AD, MCI/AD, HC/MCI/AD. CDRSB: Clinical Dementia Rating Scale Sum of Boxes; ADAS: Alzheimer’s Disease Assessment Scale; VGM: Volume of Gray Matter; VWM: Volume of White Matter; SA: Surface Area; FA: Fractional Anisotropy; MD: Mean Diffusivity; RD: Radial Diffusivity; LD: Longitudinal Diffusivity; HC:Healthy Control; MCI: Mild Cognitive Impairment; AD: Alzheimer’s disease

### 3.2. Estimating Feature Importance using ERT and Permutation Test

To estimate feature importance in this study, we predicted cognitive measures under null and real hypotheses and compared the results (**Figure 2**). The Aβ biomarker was used as the best predictor to predict MMSE scores in both hypotheses. Using 500 permutation tests for each feature importance value, the results were averaged across all subjects and reported as mean and standard deviation. Note that feature importance values calculated for the real hypothesis were averaged over 500 test split iterations based on the real value of cognitive scores. We normalized and sorted the importance of each feature across all brain regions to facilitate comparisons. As shown in **Figure 2A**, normalized importance (NI) features for the HC/MCI combination were around one for almost all brain regions. ERT algorithm assigned similar importance to brain regions as Aβ biomarkers under the null hypothesis. The real predicted values in all brain regions were also around one, showing a non-significant NI for the association between Aβ and MMSE scores. In contrast, for the HC/AD and MCI/AD combinations, the ERT algorithm assigned superior and significant feature importances (larger than 1.2) to several brain regions (between 30 and 40).

**Figure 2.**
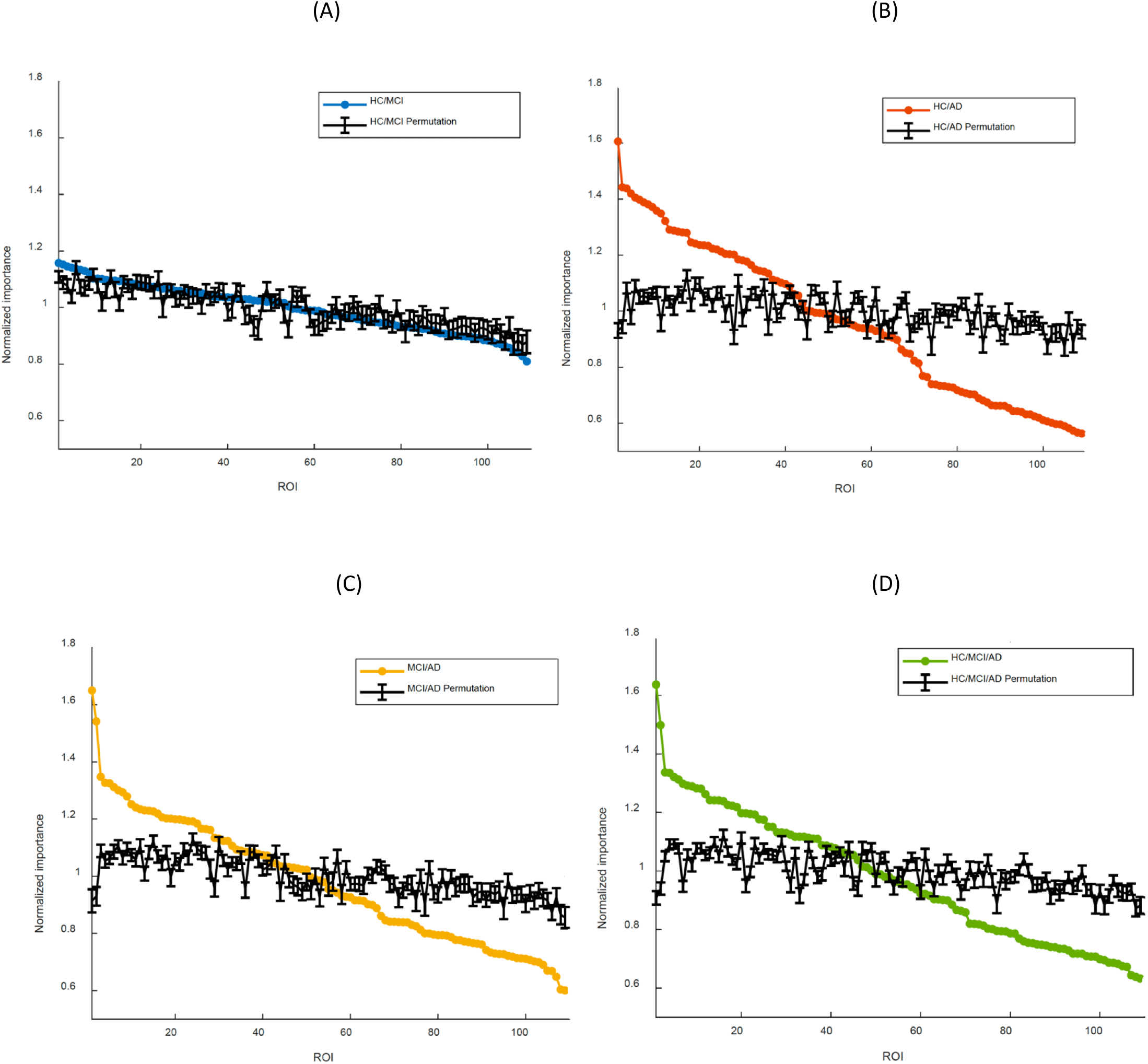
Permutation test on MMSE prediction based on Aβ biomarker in four different combinations: A) HC/MCI, B) HC/AD, C) MCI/AD, and D) HC/MCI/AD. The x-axis shows the 109 cortical and subcortical brain regions considered in the prediction of the MMSE score based on Aβ. The y-axis represents the normalized importance value for each combination of group and a random permutation. Larger values of normalized importance in brain regions indicate a higher association between that region’s feature and the MMSE score.

### 3.3. Identifying Significant Biomarkers in AD Development

As part of this study, we investigated the use of multimodal imaging biomarkers to monitor the complex interrelated mechanisms underlying AD development. The primary objective of **Figure 3-4** is to identify the most significant biomarkers across four different prediction combinations. **Figure 3A-I** compares the NI feature for MMSE prediction using nine different biomarkers and four different subject combinations. The VGM, ATH, and VWM predictors demonstrated several significant features that contributed to MMSE score prediction based on the HC/MCI combination. In contrast, the Aβ biomarker did not detect significant features in **Figure 3-I**. **Figure 3-B** revealed approximately ten features with a significant association with MMSE based on ATH. Two features observed in **Figure 3-C** were of greater importance and exhibited a strong link between the MMSE score and these two features. The presence of the AD group in the other three combinations increased the likelihood of identifying important brain regions. The Aβ biomarker showed significant effects across 40 brain regions when considering the maximum permutation NI values in all figures (Figure 3-I). **Figures 3-A, 3-B, and 3-C** with **Figure 3-I** showed that despite more features being affected by Aβ, the sMRI biomarkers VGM, VWM, and especially ATH achieved superior NI values (NI > 1.75). From **Figure 3-D**, it is evident that there are almost no SA features associated with the MMSE score; the DTI biomarkers indicate the NI values improved in three combinations of the AD group that showed at least significant NI in five features.

**Figure 3.**
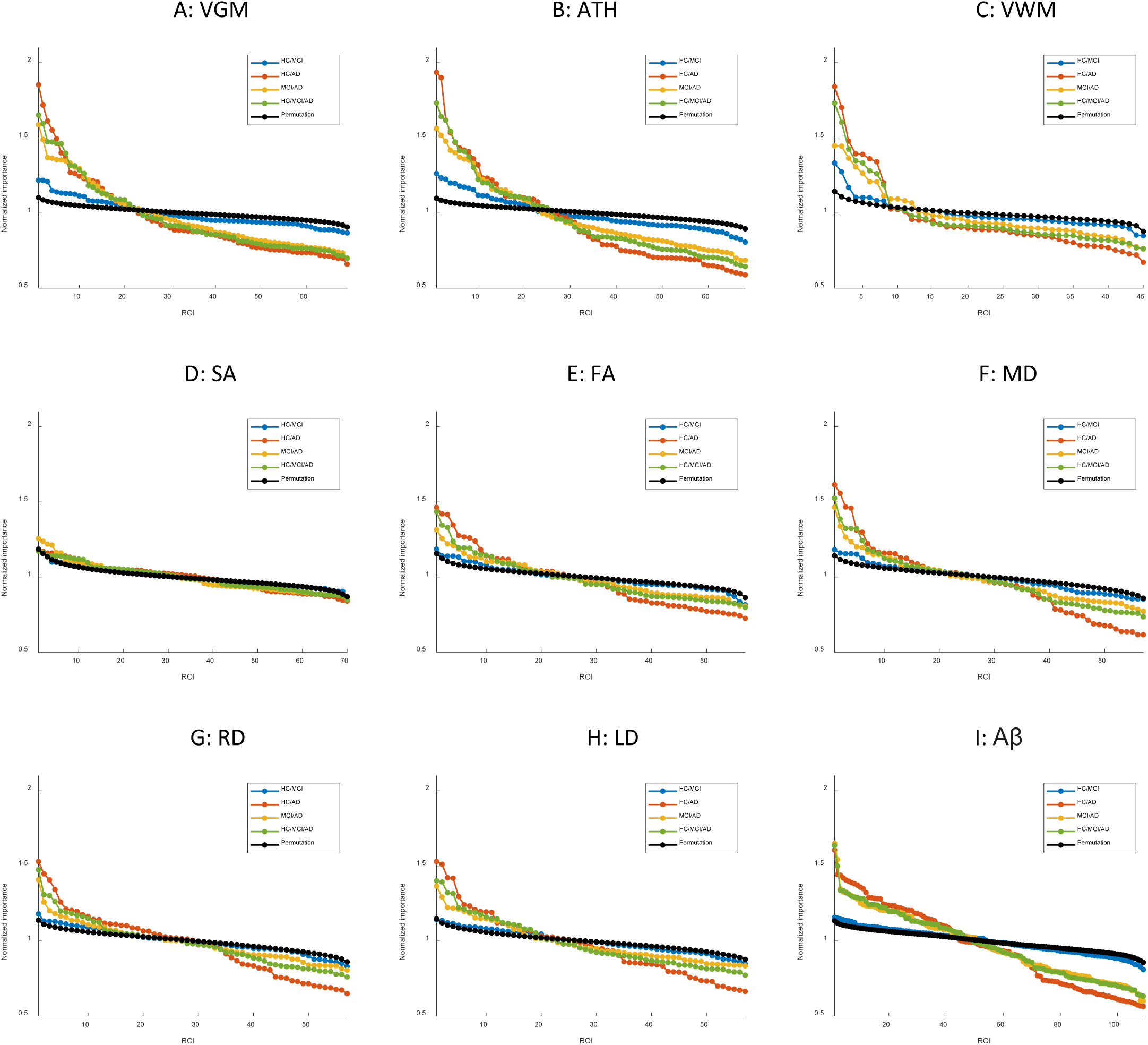
Feature importance for the prediction of MMSE score based on nine different biomarkers: A) VGM, B) ATH, C) VWM, D) SA, E) FA, F) MD, G) RD, H) LD, and I) Aβ. The x-axis shows the brain regions considered in the prediction based on each biomarker. The y-axis represents the normalized importance value for four different combinations of groups and a random permutation. Larger values of normalized importance in brain regions indicate a higher association between that region’s feature and the MMSE score.

**Figure 4.**
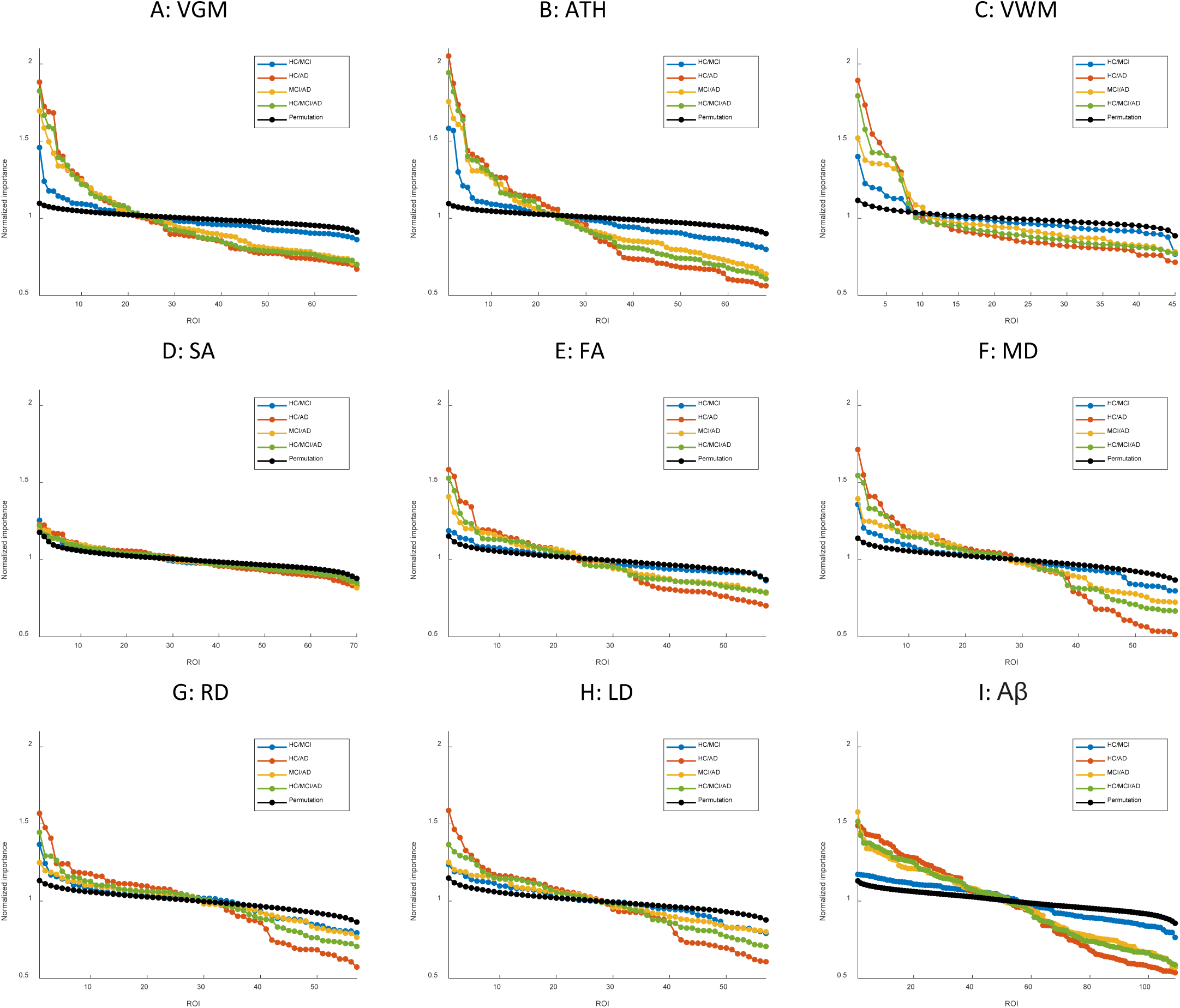
Feature importance for the prediction of ADAS score based on nine different biomarkers: A) VGM, B) ATH, C) VWM, D) SA, E) FA, F) MD, G) RD, H) LD, and I) Aβ. The x-axis shows the brain regions considered in the prediction based on each biomarker. The y-axis represents the normalized importance value for four different combinations of groups and a random permutation. Larger values of normalized importance in brain regions indicate a higher association between that region’s feature and the ADAS score.

**Figure S1** in the supplementary material illustrates the NI values in nine types of biomarkers to predict CDRSB. CDRSB prediction converges almost to the same conclusion as MMSE score prediction (shown in **Figure 3**), but the presence of the AD group in combination achieved higher NI compared to MMSE score prediction (**Figure S1-A** and **Figure S1-B**).

The NI results for ADAS prediction are shown in **Figure 4**. The NI prediction of the HC/MCI group combination provided the most interesting results in this figure. Almost all biomarkers were able to find at least one significant feature, not only in the combinations containing AD but also in the HC/MCI combination. As a result, several features were significantly affected by all nine neuroimaging biomarkers. In the HC/MCI combination, ATH was the best predictor of ADAS score, and two brain regions had NIs over 1.5. Among DTI biomarkers, MD and then RD were the most effective biomarkers. In the MCI/AD, HC/AD, and HC/MCI/AD combinations that had the AD group present, approximately 45, 25, and 15 features, respectively, contributed to the prediction of Aβ, ATH, and MD.

### 3.4. Most Discriminatory Brain Regions in All Combinations

Based on our prediction model, we observed several associations between ADAS and neuroimaging biomarkers using four different combinations. Aβ, ATH, and MD were the most important neuroimaging biomarkers for predicting cognitive scores based on PET, sMRI, and DTI, respectively. **Figures 5-7** depict the NI values of these biomarkers based on ADAS prediction and visualize them to the brain using a heat map. In the supplementary materials section, we also present the NI values of these three best biomarkers for MMSE and CDRSB prediction (**Figures S2-S7**). To illustrate the NI gradual increase, we sorted them from small to large differences between NI values in each combination, starting from HC/MCI to MCI/AD, then HC/MCI/AD, and lastly, HC/AD. We highlighted the NI values with the strongest contribution in red and yellow colors for all four combinations, and the non-significant values are represented with dark blue color.

**Figure 5.**
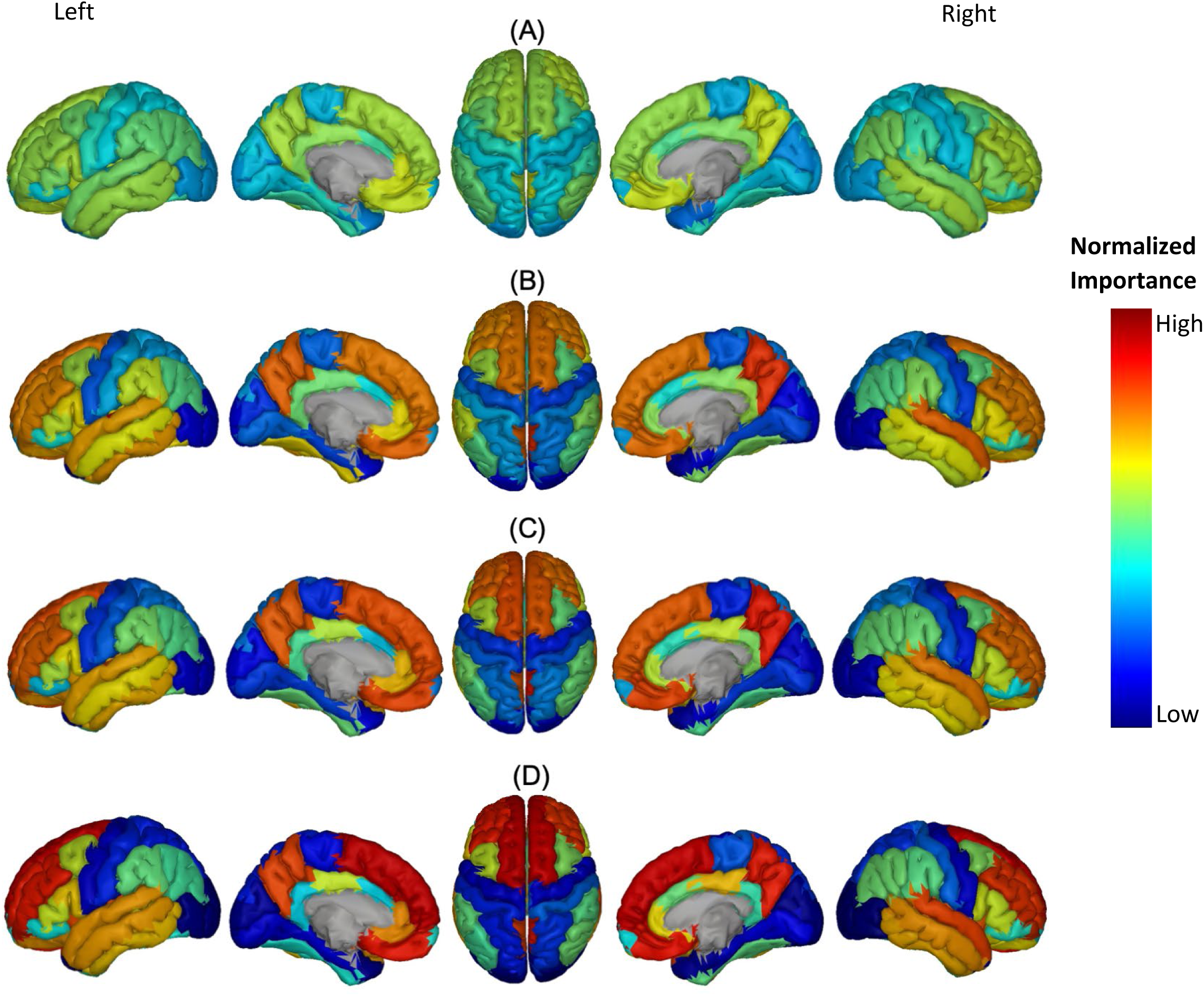
Feature importance of Aβ in cortical brain regions for the prediction of ADAS score in four combinations of groups: A) HC/MCI, B) HC/AD, C) MCI/AD, and D) HC/MCI/AD. The groups of subjects show small to large differences in the cognitive scores from normal aging to AD, from top to bottom. The color map is based on the normalized feature importance values.

The NI of cortical brain regions used in Aβ biomarkers is illustrated in **Figure 5**. The top-ranked brain regions, based on assumed thresholds (mean plus two standard deviations of the permutation test values), include: right and left transverse temporal gyrus, and right precuneus cortex in the MCI/AD combination; right transverse temporal gyrus, right precuneus cortex in the HC/MCI/AD combination; and right and left superior frontal gyrus, right and left transverse temporal gyrus, and right precuneus cortex in the HC/AD combination. Notably, the right transverse temporal gyrus and right precuneus cortex consistently showed high NI throughout three combinations with the presence of the AD group. The NI value of each feature in the HC/MCI combination was below the assumed threshold.

**Figure 6** shows the most discriminatory brain regions according to ATH. The top-ranking brain regions for the HC/MCI combination were the right and left entorhinal cortex and the right fusiform cortex. The other three combinations (MCI/AD, HC/MCI/AD, and HC/AD) had the same top-ranking regions: left and right entorhinal cortex, left inferior temporal gyrus, and left middle temporal gyrus. In combinations from one through four (1:HC/MCI, 2:MCI/AD, 3:HC/MCI/AD, and 4:HC/AD), the NI values of the left and right entorhinal cortex gradually increased, revealing the significant progression of these two regions based on ATH.

**Figure 6.**
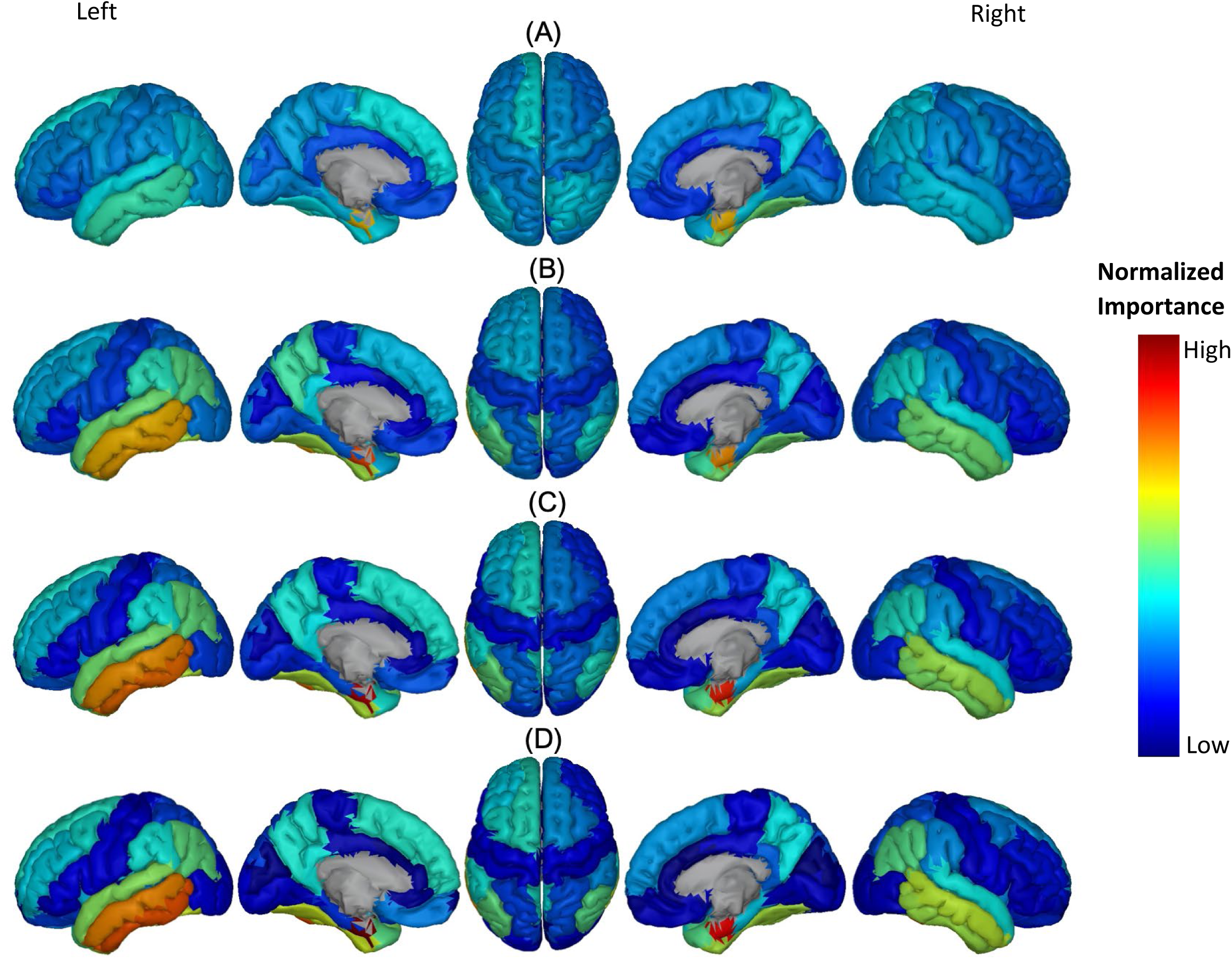
Feature importance of ATH in cortical brain regions for the prediction of ADAS score in four combinations of groups: A) HC/MCI, B) HC/AD, C) MCI/AD, and D) HC/MCI/AD. The groups of subjects show small to large differences in the cognitive scores from normal aging to AD, from top to bottom. The color map is based on the normalized feature importance values.

**Figure 7.**
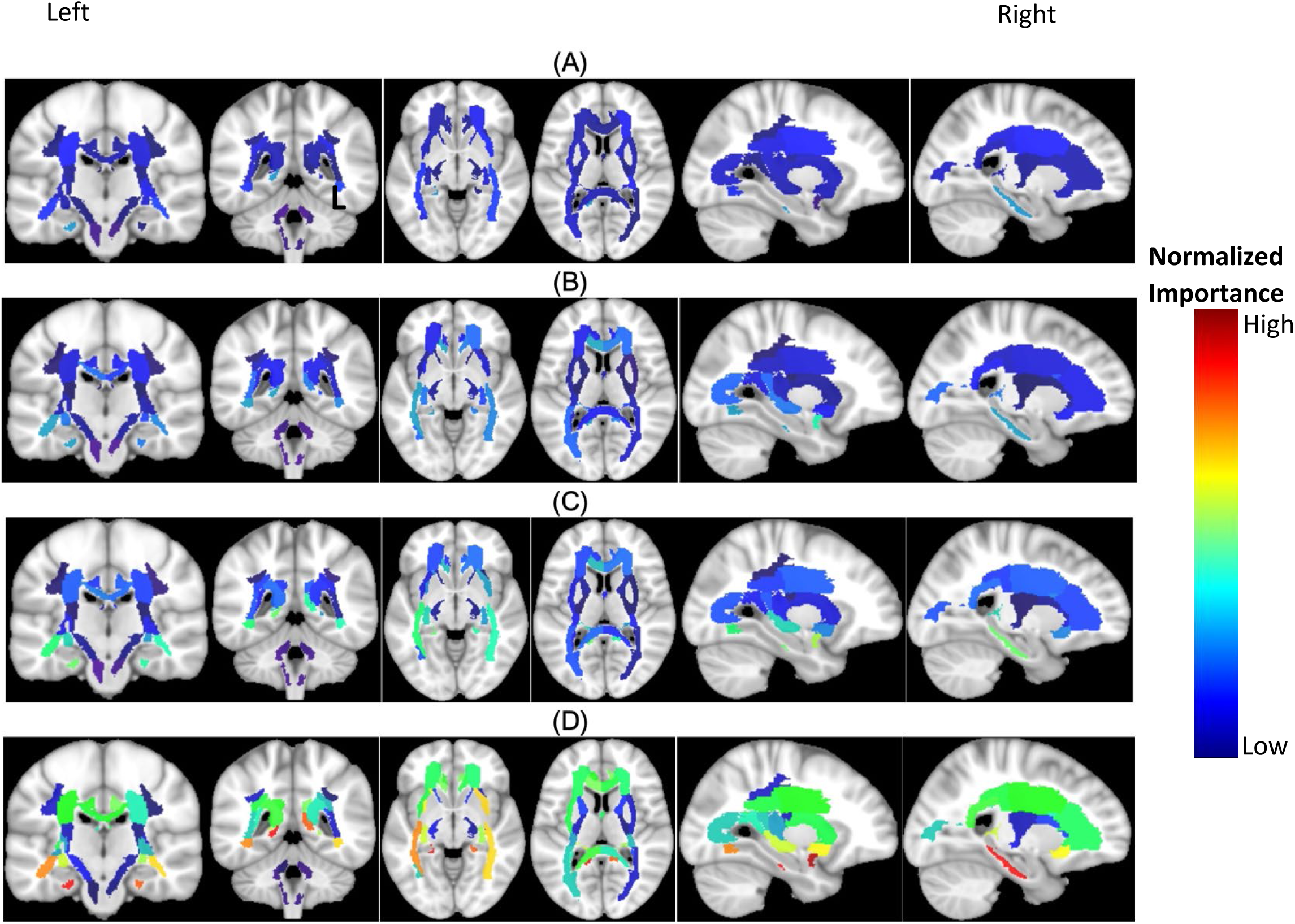
Feature importance of MD in cortical brain regions for the prediction of ADAS score in four combinations of groups: A) HC/MCI, B) HC/AD, C) MCI/AD, and D) HC/MCI/AD. The groups of subjects show small to large differences in the cognitive scores from normal aging to AD, from top to bottom. The color map is based on the normalized feature importance values.

According to previous results and as shown in **Figure 7**, MD was the most discriminatory biomarker in DTI neuroimaging. In each combination, the top regions were as follows: left cingulum and right sagittal stratum in the HC/MCI combination; left uncinate fasciculus in the MCI/AD combination; left uncinate fasciculus and left cingulum) in the HC/MCI/AD combination; and left uncinate fasciculus and left cingulum) in the HC/AD combination.

After careful consideration, we chose to focus on the HC/MCI/AD group, which encompassed all three subject groups and identified the three most effective biomarkers for each neuroimaging modality (Aβ, ATH, and MD). We used these biomarkers to demonstrate a consensus on the top features for predicting cognitive scores, as illustrated in **Figure 8**. Based on each biomarker and prediction task, we identified four overlap states: no consensus, one consensus on the cognitive score, two consensus on cognitive scores, and consensus on all three cognitive score predictions. Despite differences in biomarkers, we identified six brain regions that consistently emerged as the top regions for predicting all three cognitive scores: the right transverse temporal region for Aβ, the left and right entorhinal cortex, the left inferior temporal gyrus, the left middle temporal gyrus for ATH, and the left uncinate fasciculus for MD.

**Figure 8.**
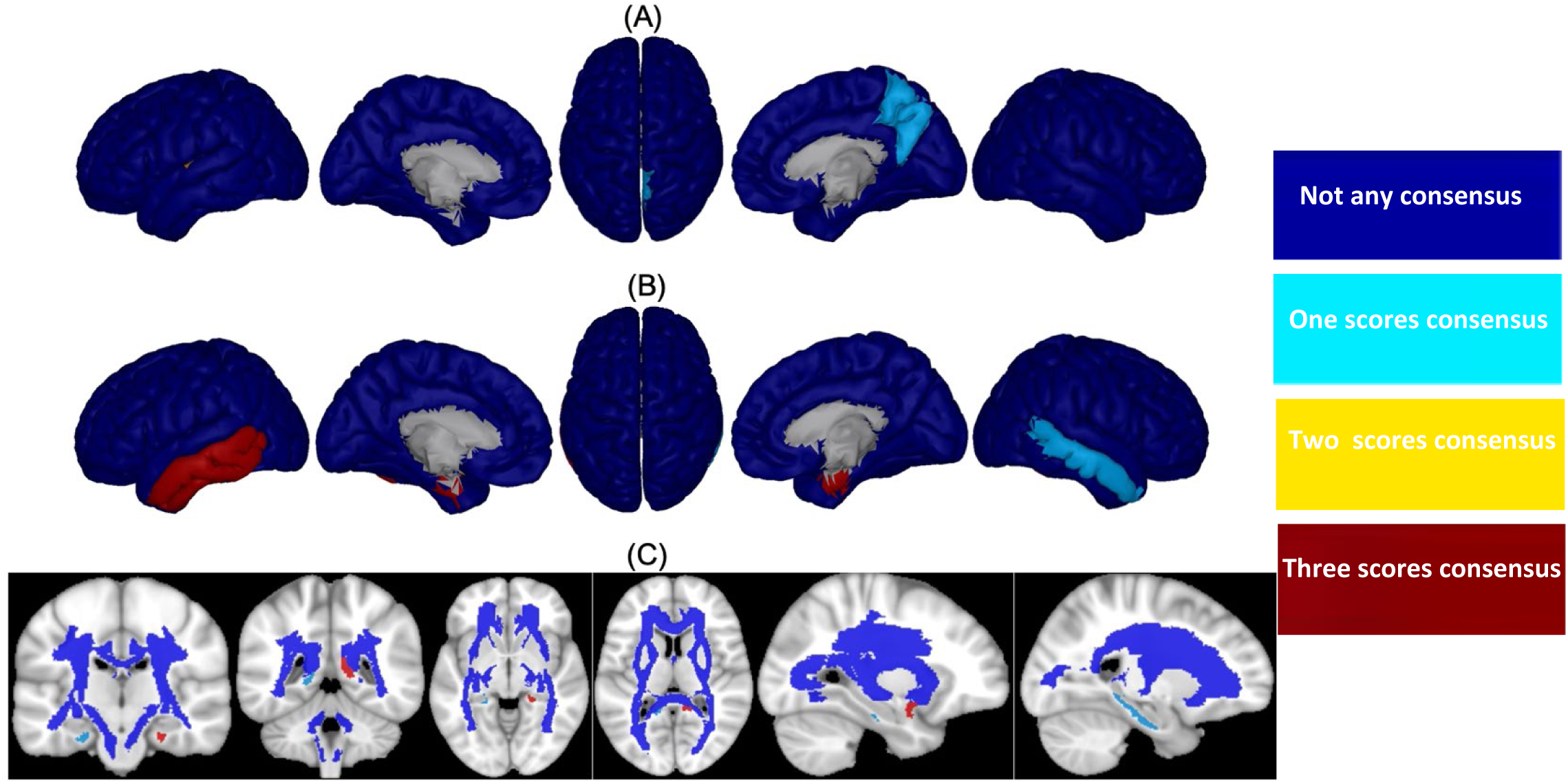
Consensus top-ranked brain regions between three cognitive tests prediction (MMSE, CDRSB, and ADAS) in HC/MCI/AD combination and based on (A) Aβ, (B) ATH, and (C) MD. The color map is calculated based on consensus between feature importance.

## 4. Discussion

AD is a complex neurological condition that affects various brain regions and cognitive functions. To better understand the link between cognitive decline and brain changes in AD, it is crucial to identify and monitor effective biomarkers and their associated brain regions. In this study, we present a framework based on ERT to identify the brain regions that are linked to cognitive abilities. The main objectives of this study are to identify the most effective biomarkers associated with cognitive scores and to tie these biomarkers to specific brain regions. Our findings reveal that Aβ outperforms all other biomarkers regarding prediction performance (MSE), but it does not appear to be associated with cognitive decline in the absence of an AD diagnosis. The association between cognitive decline and Aβ starts at the late onset of the disease. Our second finding suggests that volumetric measures, such as ATH, are strongly associated with cognitive scores, not only in early stages (combination HC/MCI) but also in late/symptomatic stages (combination MCI/AD). Additionally, we found that ADAS is associated with almost all neuroimaging biomarkers in this study and in all combinations of subjects, regardless of their symptomatology. Thus, ADAS appears to be a cognitive test that can track brain changes based on neuroimaging biomarkers throughout the early/asymptomatic to late/symptomatic phases of the disease. Lastly, our study demonstrates that the ERT technique can capture critical brain regions that have strong associations with all three cognitive scores throughout the early to late stages of the disease. These regions include (a) right transverse temporal (Aβ); (b) left and right entorhinal cortex, left inferior temporal gyrus and left middle temporal gyrus (ATH); and (c) left uncinate fasciculus (MD). Overall, our study highlights the importance of identifying effective biomarkers and their associated brain regions in AD research.

Consensus in machine learning-based AD studies has demonstrated that combining information from different neuroimaging modalities can improve classification and regression performances [15, 44–46]. Our study also supports the notion that biomarkers from different neuroimaging techniques are complementary and can offer a better understanding of AD than using each biomarker or technique alone [18, 47–50]. Although combining different feature domains may enhance the performance of machine learning methods, it may not fully utilize the complementary information present in each biomarker, and it remains challenging to understand the contribution of each feature in modality and link the feature to specific brain regions. Therefore, we aimed to separately utilize multiple neuroimaging biomarkers to predict cognitive scores in AD to handle the discrepancy between feature domains and gain new insights into the complex changes in the brain associated with AD.

Our results showed that PET with Aβ performed consistently better than the other eight biomarkers in predicting cognitive scores based on MSE. Previous research has shown that even healthy older adults with high amyloid burden have lower cognitive performance, and high levels of Aβ are strongly related to progressive cognitive decline, particularly in episodic memory and executive function [51–53]. Longitudinal studies have shown that levels of Aβ strongly related to progressive cognitive decline, and mostly affected episodic memory and executive function [52, 53]. Despite Aβ performing well in predicting cognitive scores, the calculated NI was most significant in late/symptomatic AD stages. On the other hand, volumetric measures based on sMRI, particularly ATH, showed a relatively higher NI from early/asymptomatic to late/symptomatic AD stages in comparison with Aβ. Even among DTI biomarkers, MD indicated that WM damage is strongly associated with cognitive decline [54]. These findings suggest that exploring the differences between feature domains could provide novel insights into the intricate mechanisms of AD, rather than solely focusing on improving machine learning performance by using the best modality or integrating multimodal features.

Although each biomarker and neuroimaging technique can provide valuable insights into AD, their direct and indirect relationships with each other remain unclear, which raises questions about their individual independent contributions to cognitive decline. Moreover, the lack of clarity surrounding the simultaneous and delayed relationships between these biomarkers plays a critical role in understanding the heterogeneity of AD, which remains an unresolved challenge. This ambiguity is reflected in conflicting reports regarding the relationship between the best biomarkers in our study: Aβ and ATH. Some studies suggest that an increase in Aβ deposition is linked with neurodegeneration, such as cortical thinning and/or lower volume [63–55], while others report the opposite, where higher Aβ deposition is associated with cortical thickening and/or increased volume [64–67]. Fortea et al. [68] examined the association between Aβ values and cortical thickness in a group of cognitively preserved individuals and found a complex and nonlinear (inverted-U shaped) relationship between Aβ values and cortical thickness in various brain regions. They reported that changes in cortical thickness in regions like temporoparietal regions and precuneus were linked to intermediate Aβ values that may precede cortical thinning. Wirth et al. [71] investigated non-Aβ factors of neurodegeneration within AD regions in older HC adults and found that many had neurodegenerative biomarker abnormalities in AD-affected brain regions, despite having normal Aβ levels. This evidence suggests that neurodegenerative patterns similar to AD can also develop through non-Aβ pathways and affect cognition in older adults without Aβ burden [69, 70].

Lastly, in order to better understand the relationship between cognitive scores and biomarkers, it is essential not only to determine the most effective biomarker for each neuroimaging modality, along with their respective associations with cognitive scores, but also to identify the most significant brain regions for each biomarker and modality. The ERT framework utilized in this study generated a set of features (i.e. brain regions) that were weighted and ranked based on their predictive power for cognitive scores. Prior research findings align with our results, suggesting that cognitive decline is linked to pathologies and atrophy in the temporal lobe of the brain [72]. Specifically, critical biomarkers such as the transverse temporal gyrus and precuneus cortex have been identified in the cognitive decline associated with Aβ [77]. Additionally, the cortical thickness of the entorhinal cortex has been independently and additively associated with declining memory, while different temporal regions have been identified as critical biomarkers in AD-related memory decline [78, 79]. Finally, our results are consistent with previous research that has demonstrated a significant negative association between cognitive scores and white matter integrity in the cingulum and the uncinate fasciculus [80, 81].

AD is a complex and multifaceted illness that impacts the brain and is linked to cognitive deterioration. The symptoms of AD can manifest differently, as can the underlying biological transformations in the brain. Additionally, there are several subtypes of AD that vary in the distribution of abnormal pathologies and patterns of brain changes. Employing an interdisciplinary approach to AD entails drawing on diverse neuroimaging modalities to gain a more comprehensive understanding of the disease and its underlying mechanisms. Grasping the role of each biomarker in cognitive decline is vital in overcoming the heterogeneity of AD and developing more effective treatments.

## 5. Conclusion

In this study, we investigated the potential of an interdisciplinary approach to predict cognitive scores in AD by utilizing multimodal neuroimaging biomarkers. Our proposed ERT prediction model achieved this goal by identifying the most associated biomarkers, especially in the early stages of the disease, and mapping their importance to specific brain regions. Our findings revealed that Aβ, ATH, and MD biomarkers, derived from PET, sMRI, and DTI, respectively, were strongly associated with cognitive decline in AD. Among the nine biomarkers examined, ATH had the strongest association with the cognitive disorder in the HC/MCI combination (early stage of AD), while Aβ biomarkers were most effective in predicting cognitive decline in AD-stage subjects. Furthermore, we found that ADAS decline was best explained by almost all considered biomarkers, unlike MMSE and CDRSB. Our results showed that cognitive decline was primarily driven by the right transverse temporal gyrus (based on Aβ), left and right entorhinal cortex, left inferior temporal gyrus, left middle temporal gyrus, and left uncinate fasciculus. These findings highlight the importance of an interdisciplinary approach to understanding the underlying mechanisms of AD, provide insight into the heterogeneity of this disease, and may help in the development of more effective treatments.

## Abbreviation Definition

AD: Alzheimer’s Disease
Aβ: Amyloid-β
ADAS: Alzheimer’s Disease Assessment Scale-Cognitive Subscale
ATH: Average thickness
BT: Bagging tree
CDRSB: Clinical Dementia Rating Scale Sum of Boxes
DTI: Diffusion tensor imaging
ERT: Ensemble regression tree
FA: fractional anisotropy
FDG: 18F-fluorodeoxyglucose
HC: Healthy controls
LD: Longitudinal diffusivity
MD: Mean diffusivity
MCI: Mild cognitive impairment
MMSE: Mini-Mental State Examination
MSE: Mean square error
NI: Normalized importance
PET: Positron emission tomography
RD: Radial diffusivity
RF: Random forest
sMRI: Structural magnetic resonance imaging
SA: surface region
VGM: volume of gray matter
VWM: volume of white matter

## Data Availability

The data that have been used in this study is publicly available.

https://adni.loni.usc.edu/

---

**Figure S1.**
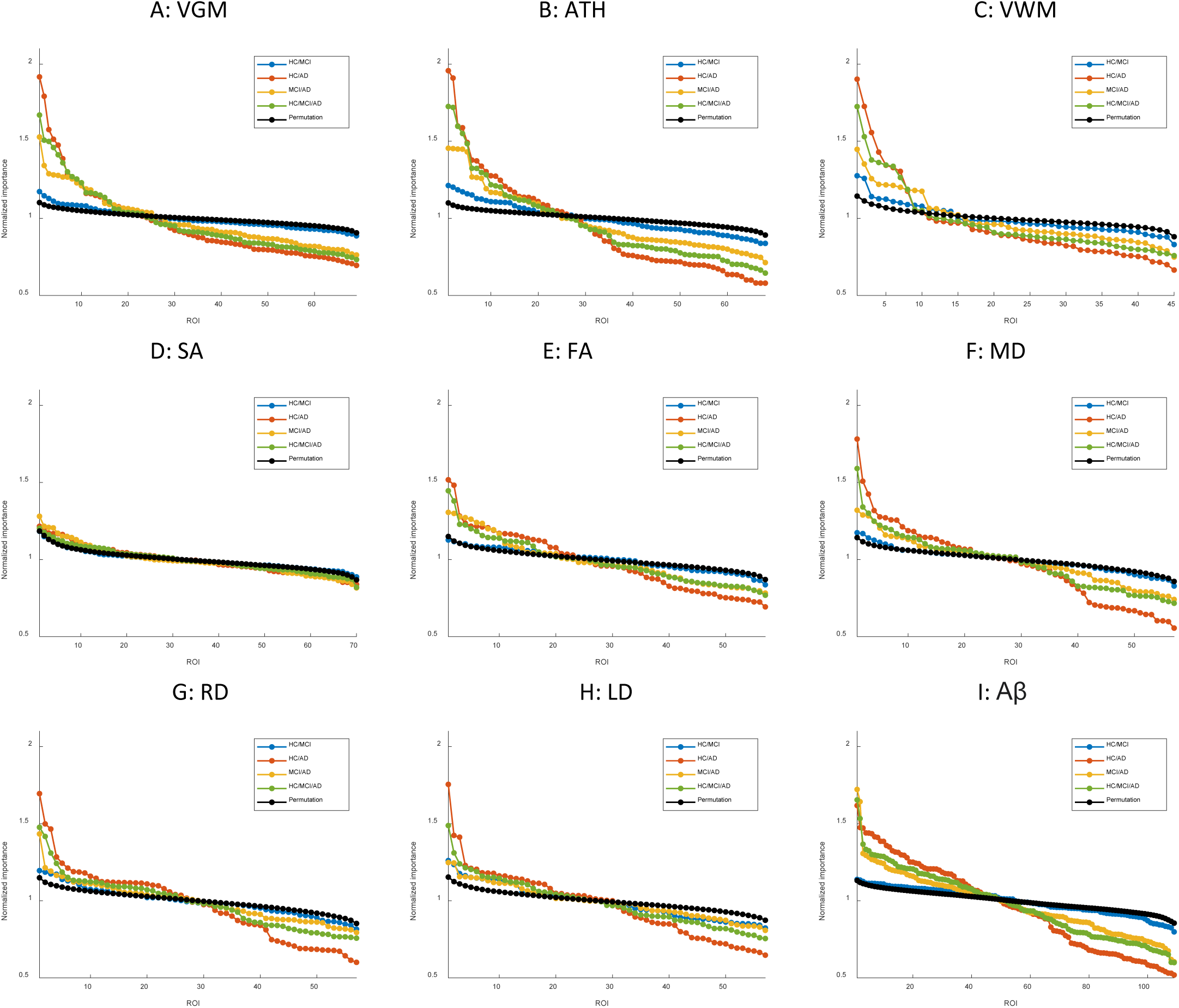
Feature importance for the prediction of CDRSB score based on nine different biomarkers: A) VGM, B) ATH, C) VWM, D) SA, E) FA, F) MD, G) RD, H) LD, and I) Aβ. The x-axis represents the brain regions considered in the prediction based on each biomarker. The y-axis represents the normalized importance value for four different combinations of groups and a random permutation. Larger values of normalized importance in brain regions show a higher association between that region’s feature and the CDRSB score.

**Figure S2.**
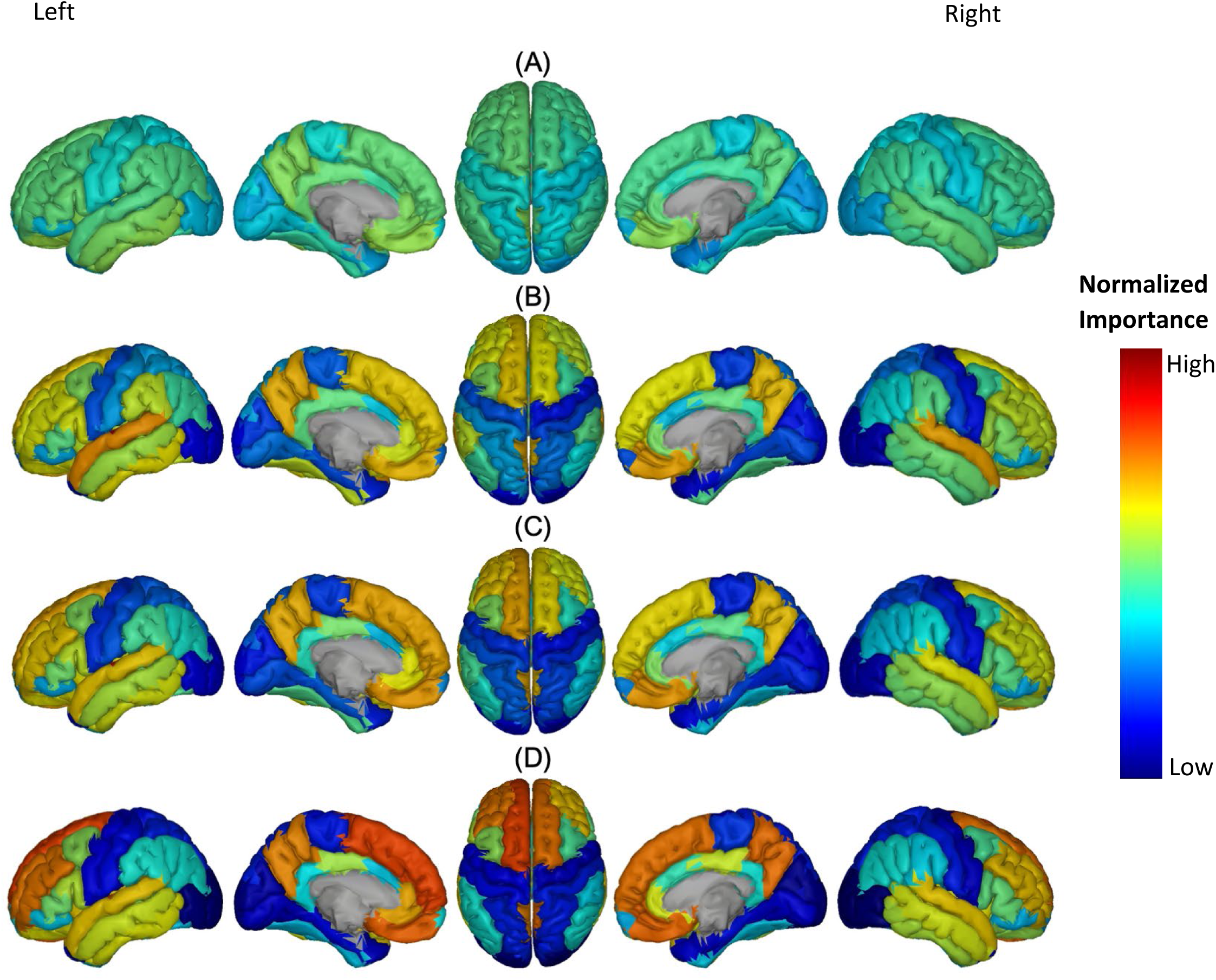
Feature importance of Aβ of the cortical brain regions for predicting the MMSE score in four different combinations of groups: (A) HC/MCI, (B) MCI/AD, (C) HC/MCI/AD, and (D) HC/AD. The groups of subjects from top to bottom show a gradual increase in cognitive decline from normal aging to AD. The color map was calculated based on normalized feature importance values, which indicate the degree of association between each cortical brain region’s feature and the MMSE score.

**Figure S3.**
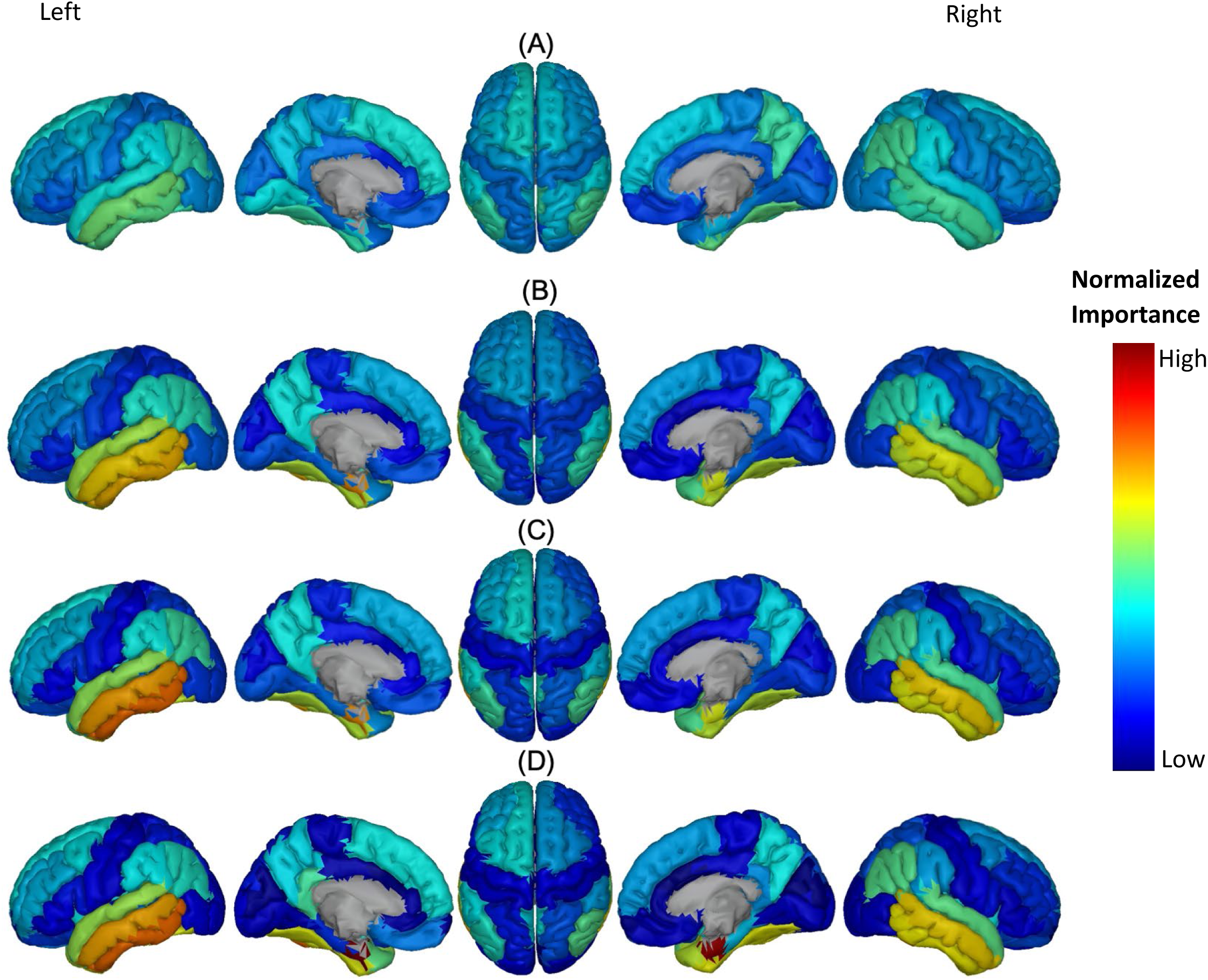
Feature importance of ATH of the cortical brain regions for the prediction of MMSE score in four combinations of groups: (A) HC/MCI, (B) MCI/AD, (C) HC/MCI/AD, and (D) HC/AD. From top to bottom, groups of subjects have small to large differences in the cognitive scores from normal aging to AD. The color map was calculated based on normalized feature importance values.

**Figure S4.**
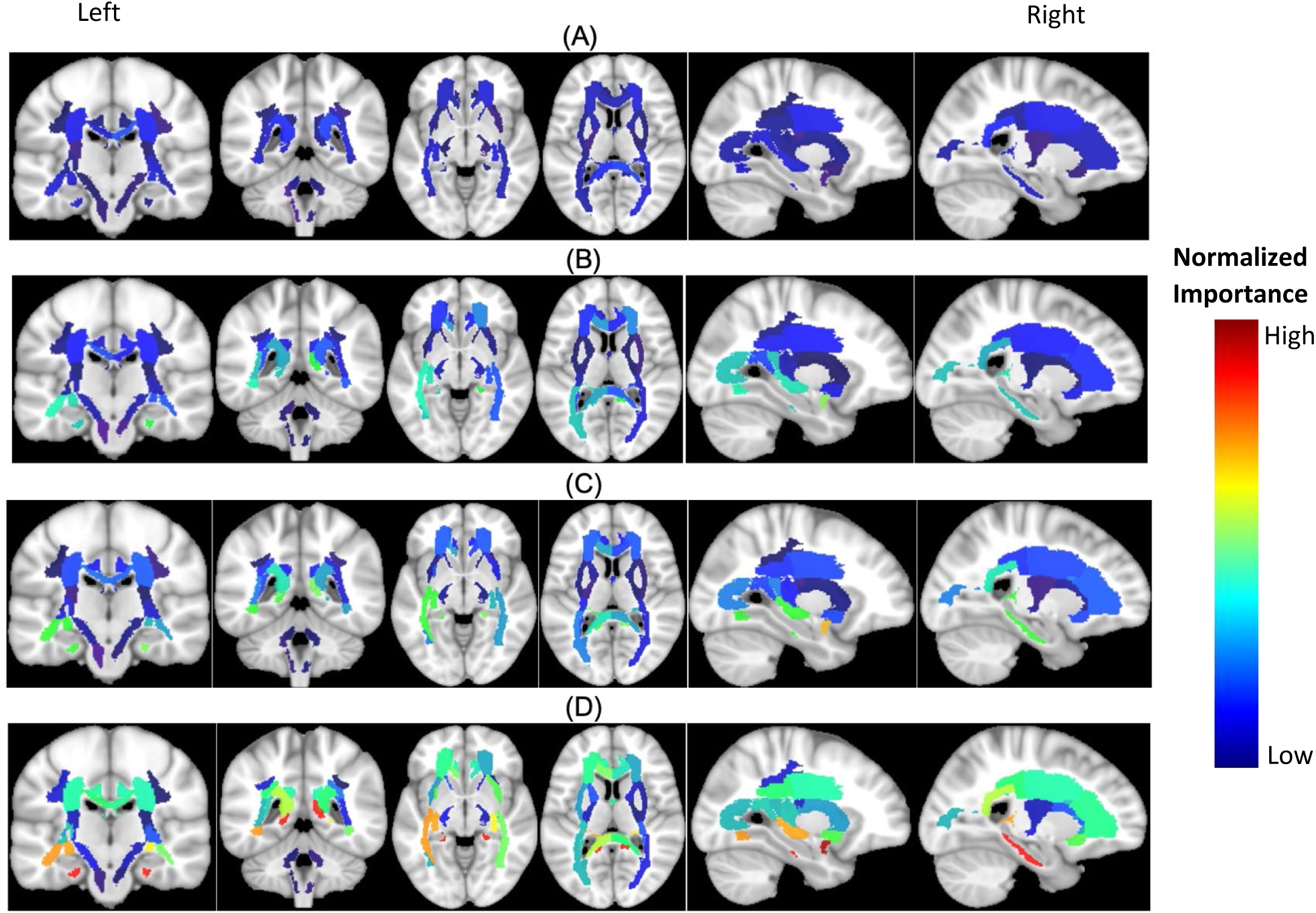
Feature importance of MD of the cortical brain regions for the prediction of MMSE score in four combinations of groups: (A) HC/MCI, (B) MCI/AD, (C) HC/MCI/AD, and (D) HC/AD. From top to bottom, groups of subjects have small to large differences in the cognitive scores from normal aging to AD. The color map was calculated based on normalized feature importance values.

**Figure S5.**
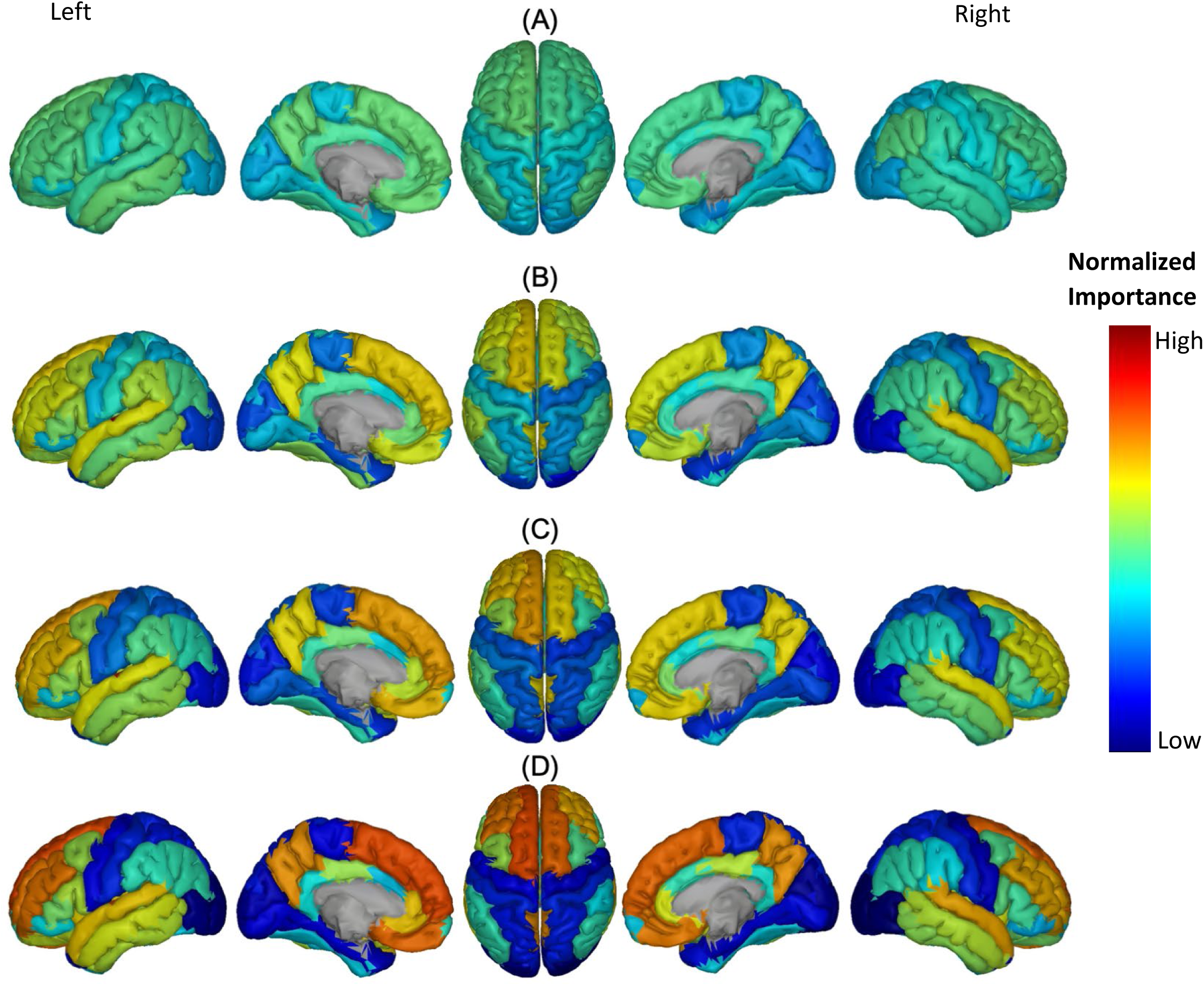
Feature importance of Aβ of the cortical brain regions for the prediction of CDRSB score in four combinations of groups: (A) HC/MCI, (B) MCI/AD, (C) HC/MCI/AD, and (D) HC/AD. From top to bottom, groups of subjects have small to large differences in the cognitive scores from normal aging to AD. The color map was calculated based on normalized feature importance values.

**Figure S6.**
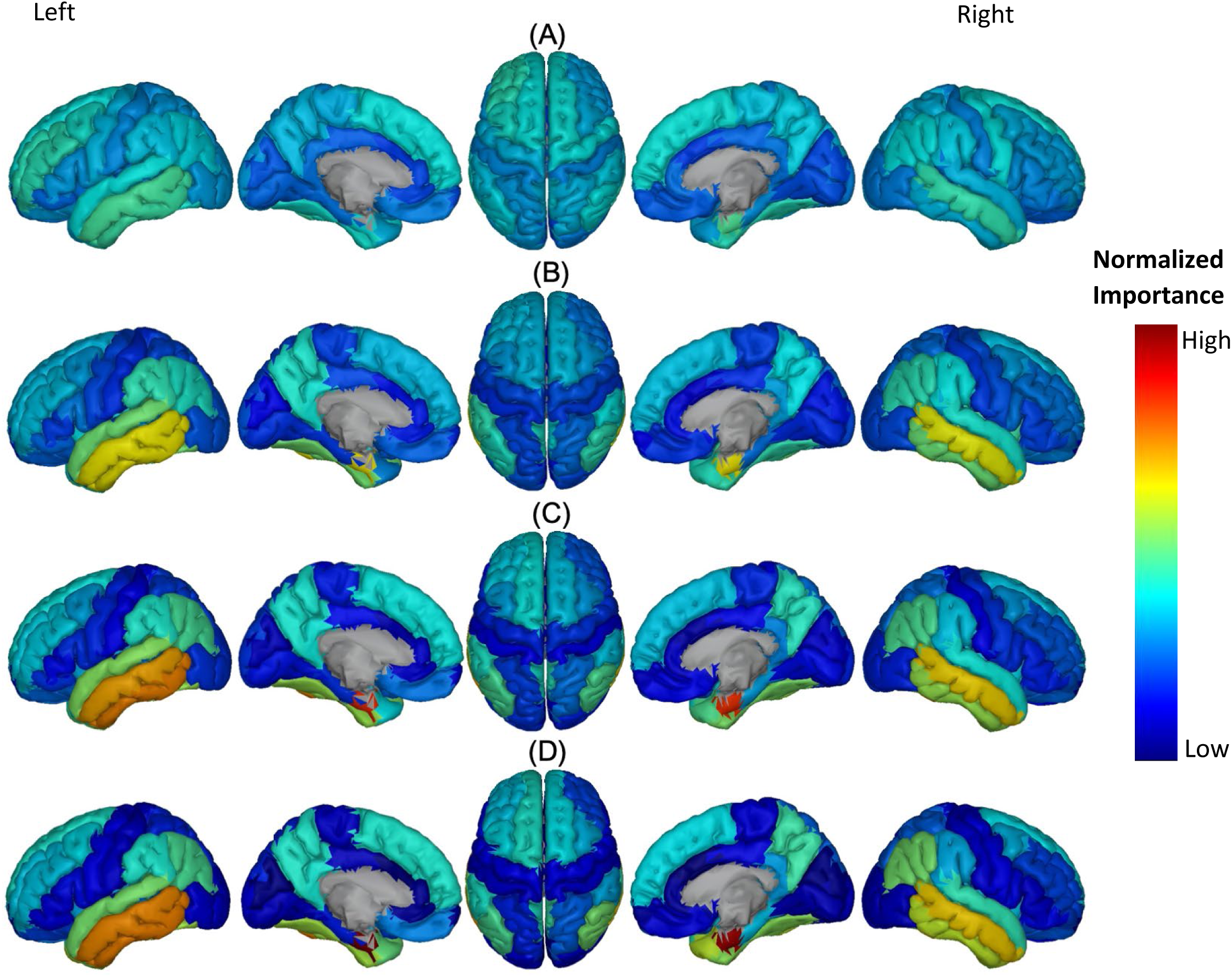
Feature importance of ATH of the cortical brain regions for the prediction of CDRSB score in four combinations of groups: (A) HC/MCI, (B) MCI/AD, (C) HC/MCI/AD, and (D) HC/AD. From top to bottom, groups of subjects have small to large differences in the cognitive scores from normal aging to AD. The color map was calculated based on normalized feature importance values.

**Figure S7.**
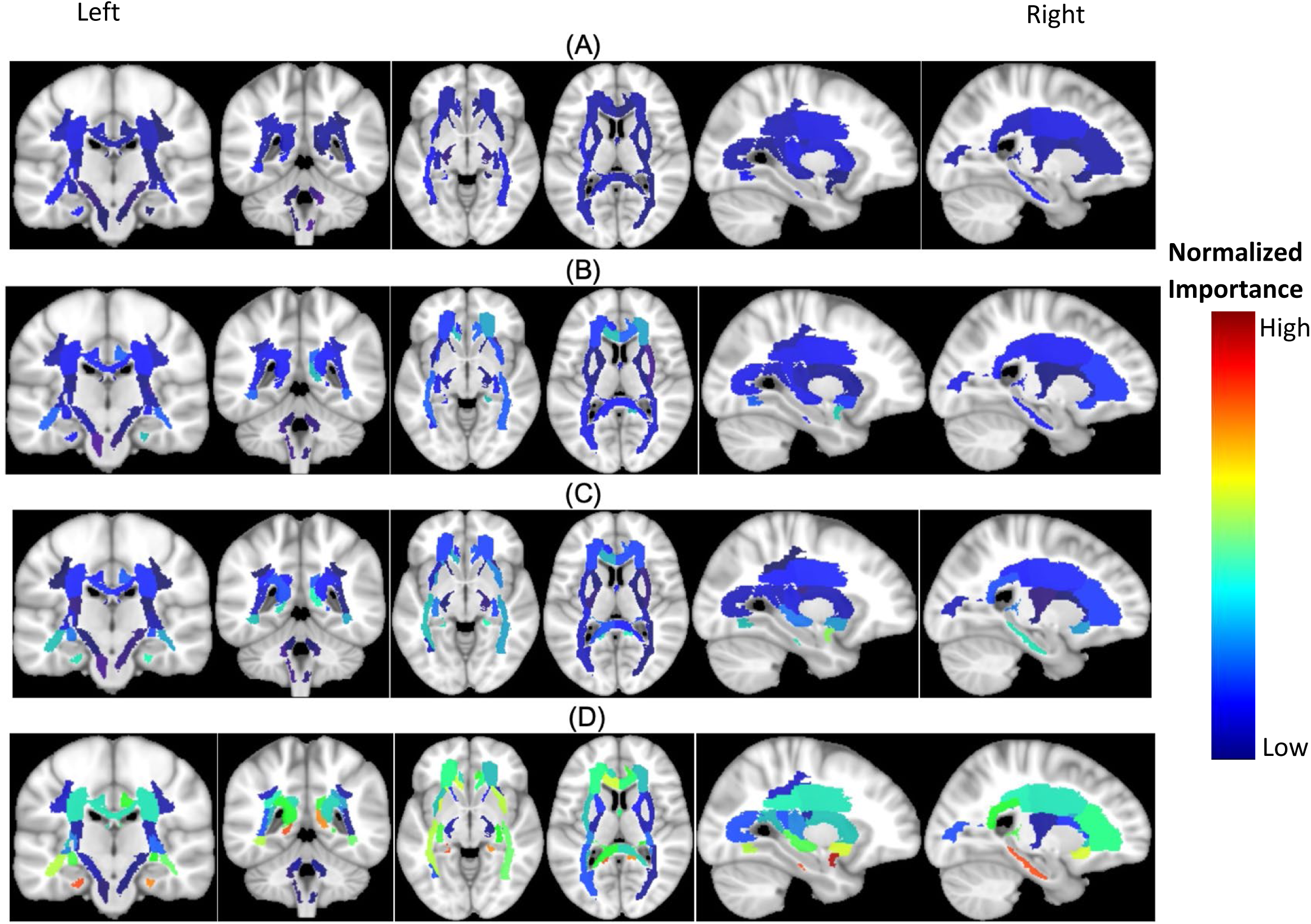
Feature importance of MD of the cortical brain regions for the prediction of CDRSB score in four combinations of groups: (A) HC/MCI, (B) MCI/AD, (C) HC/MCI/AD, and (D) HC/AD. From top to bottom, groups of subjects have small to large differences in the cognitive scores from normal aging to AD. The color map was calculated based on normalized feature importance values.

